# Structured Onboarding Feasibility in Community EDs

**DOI:** 10.64898/2026.02.15.26346347

**Authors:** Peter Guertin, Kelly R. Conner, Vijay Nagpal

**Affiliations:** Wake Forest University School of Medicine. Winston-Salem, NC; Wake Forest Emergency Providers. Winston-Salem, NC. Now affiliated with Hospital Corporation of America – Doctors Hospital of Augusta. Augusta, GA

**Keywords:** Advanced Practice Providers, Emergency Medicine, Transition to Practice, Workforce Retention

## Abstract

**Background:** Advanced Practice Providers (APPs), including physician assistants and nurse practitioners, represent a growing proportion of the emergency medicine workforce, including in high-acuity community emergency departments (EDs). Despite this growth, many sites lack formal onboarding structures, particularly for new graduate or inexperienced APPs transitioning to practice. Unlike postgraduate residencies and fellowships, limited literature exists on structured onboarding models outside academic settings. This study evaluated the feasibility and perceived impact of a structured onboarding program for APPs in a non-academic community ED.

**Methods:** This mixed-methods feasibility study was conducted at a single-site community ED without an existing formal onboarding process. New graduate or inexperienced APPs hired within 12 months of program implementation completed a post-intervention survey assessing satisfaction across five domains derived from a conceptual framework of human resource practices and retention. Quantitative data was collected using 5-point Likert-scale items, and qualitative data was obtained through open responses. Leadership and preceptors completed a secondary survey evaluating feasibility and perceived impact. Descriptive statistics and thematic analysis were performed.

**Results:** Four new graduate APPs (100% response rate) completed the post-implementation survey. Mean scores across domains ranged from 3.33 to 5.00, with highest ratings observed in supervisor support (mean = 5.00), employee engagement (4.33), and alternative training via online modules (4.67). Qualitative themes included clear communication of expectations, value of asynchronous educational modules, and strong mentorship support. Fifteen leaders and preceptors reported that although the program required additional effort, it improved tracking of APP progress, preparedness for transition to practice (4.67), and was perceived as worthwhile to reduce attrition.

**Conclusions:** A structured onboarding program for new graduate APPs in a community ED was feasible, well accepted, and perceived to support transition to practice. These findings support the need for further study of structured onboarding as a scalable strategy to enhance preparedness, engagement, and potential retention in high-acuity clinical settings.

## Introduction

Advanced Practice Providers (APPs) including Physician Assistants and Nurse Practitioners make up a substantial portion of the workforce in high acuity specialties and subspecialties. It is estimated that from 2010 to 2017, APPs were involved with 21% of all Emergency Department (ED) visits in the United States.^1^ With the increasing number of patients being treated by APPs, there is increasing need to safely and effectively onboard them into the workforce. Two of the most important factors surrounding APP onboarding identified by healthcare leaders are retention and ensuring quality care.^2^ However, healthcare leaders face the additional challenge of bringing a growing number of new graduate or inexperienced APPs into the workforce.^3^ Developing a process for integration into departmental operations, professional development has shown to improve APP job satisfaction as well as improve APP retention in Emergency Medicine.^4^ The cost to recruit, hire and onboard an APP is widely varied, but has been reported to be as much as $250,000. With rising healthcare costs and evolving reimbursement models, retention has become an important factor to reduce overall expenditure.^5^

There are many factors that have been identified that contribute to the overall trajectory of an APP’s employment within a healthcare organization from the recruitment and hiring phase, to the integration and operational phase, to the retention and advancement phase. These factors can be influenced at the system level, service line or departmental level, and the individual level. Relevant factors to integration and retention that were identified included organizational culture, resources to support roles and integration, training opportunities, autonomy and relationship with collaborating physician, career development and advancement pathways, and perception of organizational leadership.^3^ Despite the knowledge of the factors contributing to APP success, no uniform process has been developed to increase APP satisfaction with their onboarding process and development to help decrease attrition and drive down the costs associated with it.^4^ APPs have previously identified specific training points such as improving clinical competence, training on the Electronic Health Record (EHR), and orientation to organization dynamics as factors contributing to the success of their onboarding.^6^ While there is an adequate body of literature describing the success of formal residencies and fellowship programs, there is a gap in the literature detailing formal onboarding processes.^7^ Despite a lack of literature describing a formal onboarding process, there is literature suggesting that the very existence of a formal structure on the onboarding process improves the success of an APP’s transition to practice in high acuity practice environments.^8^ Through a formal onboarding process, APPs can not only increase competency and learn proactivity but can also increase social connections and undergo mentorship which has been proven to increase retention and improves organizational culture.^9^ While a structured approach is important, there must be flexibility within the framework to ensure that there is room to mold the process to individual needs.^10^

The primary aim of this study was to assess the feasibility of a formalized onboarding process for new graduate and inexperienced APPs. The secondary aim was to assess the perception of the use of this onboarding by site leadership and preceptors.

### Methodology

This was a feasibility study utilizing convenience sampling due to the ease of access to the APPs who were undergoing onboarding at a single site community ED that previously had no formal onboarding plan and does not associate with an academic medical center or post graduate fellowship or residency program. The potential population was any new graduate or inexperienced APP hired within the 12 months following the implementation of the structured onboarding process of 8 weeks. (Supplemental Materials: Appendix A). A new graduate was defined as any APP for whom this was their first job after completion of their training, not including post graduate fellowships. Inexperienced APPs were defined as any APP who had less than 1 year of experience in Emergency Medicine, Critical Care, or other high acuity specialties.

IRB approval was sought from the Advocate Health Enterprise IRB (IRB00122626), though no ethical issues were anticipated as participants would have undergone an unstructured onboarding process otherwise and requesting evaluation of processes is a standard practice. Due to this, consent was waived. To limit potential conflicts of interest due to researchers’ supervisory relationships with participants, researchers were blinded to participant responses.

Quantitative and qualitative data was gathered from participants post intervention and at the midpoint of the intervention. For the quantitative analysis, the data was analyzed using correlation analysis to establish a baseline in satisfaction with the onboarding process. The preceptor qualitative data was used to assess baseline feasibility with the process itself. Any signals for benefit were identified even within the given limitations and then overlayed with open-ended responses from the qualitative data.

Data was gathered using a 14-question survey designed to address the most important aspects of onboarding and retention identified in the existing literature consisting of twelve 5-point Likert-scale responses from Strongly Disagree to Strongly Agree and 2 open-ended responses for qualitative data gathering (Supplemental Materials: Appendix B). This research adapted the Conceptual Framework of HR practices and employee retention to approach the problem of APP satisfaction with structured onboarding and transition to practice as well as increase retention. This framework incorporates the domains of Career Development, Training and Development, Performance Appraisal, Reward and Compensation, and Health and Safety to achieve the overall goal of retention.^12^ The framework described above identified 5 domains contributing to onboarding and retention, 2 questions were dedicated to each domain. Each question was designed to address an individual variable within that domain. The questionnaire was derived from similar existing questionnaires addressing the impacts of employee training.^18,19,20^ External validation of this survey was not sought prior to use due to the pilot nature of this study. However, to help increase strength and face validity of these survey questions, draft questions were put forth to second year PA students and refined to ensure that they were appropriately perceived. Quantitative data was gathered using questions with a 5-point ordinal Likert scale. REDCap electronic data capture software hosted by Wake Forest University School of Medicine was used to distribute the survey. Calculations occurred for each individual question as they are associated to different variables. There were two questions aimed at assessing overall satisfaction to establish a baseline.

Open-ended questions were included to allow respondents to identify perceived highlights of the intervention, shortfalls, and make suggestions for change during future iterations. The open-ended responses were audited by 3 independent reviewers. The Lead APP, the Supervising Physician, and the site Medical Director. The reviewers met to develop the code book until deductive thematic saturation was reached. They met regularly to ensure inter-rater reliability. All responses were scored based on the code book. Qualitative responses were overlayed with ordinal scale questions to help support any identified signals for benefit. The responses underwent further thematic analysis to identify any aspects of onboarding contributing to satisfaction that were not addressed in the intervention but could be included in future iterations.

Ba A 5 question 5-point Likert scale survey was distributed to the Medical Director, Supervising Physician, and any preceptor involved in greater than 6 shifts of the onboarding process (Supplemental Materials: Appendix C). REDCap software was used to distribute the survey. This equated to about 25% of the APP’s onboarding process. This number was chosen because it goes beyond the current site-specific expectation of 2-3 precepting shifts per month for various medical learners and shadowing obligations for the group.

### Participant Quantitative Results

A total of four APPs, all new graduates, completed the post-implementation survey evaluating the structured onboarding and clinical orientation process. This represented a 100% response rate among eligible participants. The participants included 3 females and 1 male, all between the ages of 20 and 30. Two of the participants underwent their physician assistant training program in Pennsylvania, one in North Carolina, and one in Tennessee. This was initial employment after board certification for all participants. Quantitative analysis focused on mean Likert-scale ratings (1 = strongly disagree to 5 = strongly agree) across five domains as well as an overall assessment which evaluated twelve total variables derived from the conceptual framework for human resource practices and retention. Descriptive statistics are presented in Table 1.

**Table 1:** Participant Quantitative Data.

| **Domain** | **Variable** | **Mean** |
| --- | --- | --- |
| **Career Development** | Career Management | 4.00 |
|  | Self-Assessment | 4.00 |
| **Training and Development** | On-the-Job Training | 3.33 |
|  | Off-the-Job (Online Modules) | 4.67 |
| **Performance Appraisal** | Communication of Expectations | 4.00 |
|  | Regular Feedback / Review | 4.33 |
| **Reward and Recognition** | Performance-Based Rewards | 4.00 |
|  | Non-Monetary Rewards | 4.00 |
| **Employee Retention** | Supervisor Support | **5.00** |
|  | Employee Engagement | 4.33 |
| **Overall Assessment** | Taught Necessary Skills | 4.33 |
|  | Performance Improved Due to Onboarding | 4.00 |

#### Domain 1: Career Development

The domain of Career Development consisted of two variables: Career Management and Self-Assessment. The mean score was 4.0 for both variables, indicating “agree” that the structured onboarding process clarified professional pathways and supported reflection on readiness for clinical practice.

#### Domain 2: Training Development

Within the Training and Development domain, On-the-Job Training received a mean score of 3.33, whereas Off-the-Job Training (online modules) scored notably higher at 4.67.

#### Domain 3: Performance Appraisal

The Performance Appraisal domain demonstrated consistently high results. The variable Communication of Expectations yielded a mean score of 4.0, while Regular Feedback and Review mean was 4.33.

#### Domain 4: Reward and Recognition

Scores within the Reward and Recognition domain were both 4.0 for Performance-Based Rewards variable and the non-monetary rewards variable, indicating that participants “agree” the onboarding process promoted fair acknowledgment of effort and non-monetary rewards such as improved work-life balance and opportunities for growth.

#### Domain 5: Employee Retention

The Employee Retention domain yielded the highest overall satisfaction ratings. Supervisor Support variable received a perfect mean score of 5.0, while Employee Engagement variable has a mean of 4.33.

#### Overall Assessment

Two global items assessed participants’ overall impressions of the onboarding program. The statement “This formal onboarding process taught me the skills I needed to perform my job” received a mean rating of 4.33, while “My performance improved due to the development and enhancement of skills required for transition to practice” scored 4.0. These statements were aimed at assessing an overall view of the program as whole.

### Participant Qualitative Results

Qualitative analysis of open-ended survey responses identified three primary themes that complemented the quantitative findings: communication about expectations and progress, value of online educational modules, and supervisor support and mentorship. Comments supporting this are summarized in Table 2.

**Table 2:** Participant Qualitative Themes.

| Theme | Description | Representative Quote |
| --- | --- | --- |
| Communication About Expectations and Progress | Participants valued clear communication of progressive performance benchmarks and workload expectations. | “The gradual increase in expectations regarding patient load, performance, and efficiency.” |
| Value of Online Modules | HIPPO EM modules were consistently identified as beneficial for reinforcing knowledge and confidence. | “The HIPPO EM modules were helpful and relevant.” |
| Supervisor Support and Mentorship | Regular one-on-one feedback from supervising physicians and APP mentors enhanced confidence and competence. | “1-on-1 feedback from the physician/APP mentor was very helpful.” |

#### Theme 1: Communication About Expectations and Progress

All participants described the value of transparent communication regarding performance expectations and progression benchmarks. This structured escalation of responsibility was perceived as a confidence-building mechanism that balanced support with growing autonomy.

#### Theme 2: Value of Online Modules

Each APP identified the HIPPO EM modules as a meaningful component of the program. Participants emphasized that the asynchronous learning format allowed reinforcement of clinical decision-making skills and efficient review of high-acuity topics. These comments corresponded with the high quantitative score (Mean Score = 4.67) for off-the-job training, further validating the integration of structured e-learning into onboarding curricula.

#### Theme 3: Supervisor Support and Mentorship

A strong mentoring relationship emerged as a central theme. Participants described the mentoring as highly beneficial in promoting professional growth. This theme aligned closely with the perfect mean rating of 5.0 for supervisor support and underscored the program’s emphasis on individualized guidance.

### Leadership and Preceptor Data

Fifteen site leaders and preceptors completed the secondary survey evaluating the feasibility and perceived impact of the structured onboarding process. Preceptors included 6 physicians and 9 APPs, 8 males and 7 females, experience ranging from 2 years to greater than 20 years in Emergency Medicine. Descriptive statistics are presented in Table 3. Respondents moderately agreed that the new onboarding process required additional time and effort compared with the previous unstructured model (Mean score = 3.13). However, they strongly agreed that the additional investment was worthwhile to reduce attrition (Mean score = 4.20) and that the new process improved the ability to track APP progress (Mean score = 4.00). Participants also agreed that the increased onboarding time did not impose undue stress on site resources (Mean score = 4.20) and, most notably, that the process improved new hire preparedness for transition to practice (Mean score = 4.67). These findings suggest widespread support among leadership and preceptors for continuing the structured onboarding model despite increased effort requirements.

**Table 3:** Leadership and Preceptor Survey.

| **Survey Item** | **Mean** |
| --- | --- |
| The new structured onboarding process required more time and effort than the previous unstructured onboarding process. | 3.13 |
| Any additional time and effort spent on the new onboarding process was worth it to reduce attrition. | 4.20 |
| It was easier to track the APP's progress through the onboarding process after the implementation of the structured onboarding process. | 4.00 |
| The increased onboarding time did not put undue stress on the site's resources. | 4.20 |
| The new onboarding process improved the new-hire APP's preparedness for transition to practice. | **4.67** |

## Discussion

This was a feasibility study designed to examine the implementation of a structured onboarding program for newly graduated APPs in a non-academic community emergency department. The findings showed that the onboarding model was feasible to implement, well-received by new hire APPs, and positively evaluated by site leadership and current providers. Quantitative and qualitative data both demonstrated that structured onboarding supported new APPs’ transition to practice by improving perceived readiness, clarifying expectations, and enhancing supervisory support. These findings contribute to the existing literature on transition to practice processes for APPs and underscores the importance of structured orientation and mentorship within high-acuity care environments.^2,4^

The strongest evidence of this program’s impact was in the domains of supervisor support, performance appraisal, and online training resources. In the Employee Retention domain, Supervisor Support received a perfect mean score, which was reinforced by qualitative comments describing the value of mentorship. The emphasis on direct access to physician and APP mentors seems to have been the key factor in improving participants’ confidence and engagement in the process. Their scores surrounding engagement also support that structured onboarding strengthens early professional connection and commitment, both of which are associated with improved retention outcomes.^4^

Performance Appraisal also showed strong results amongst the new hire APPs. Participants reported clear expectations and meaningful feedback. Qualitative feedback specifically cited the usefulness of “gradual increases in expectations” and the predictability of weekly check-ins. These findings are consistent with evidence that structured feedback process promoted early clinical competence and helped new graduate APPs integrate into the healthcare team. In EM settings, transparent performance expectations are essential for successful onboarding.^6^

Training and Development findings revealed a distinction between on-the-job training (mean 3.33) and off-the-job training via HIPPO EM modules (mean 4.67). New hire APPs consistently cited the online modules as valuable, relevant, and confidence-building. This suggests that structured e-learning served as an important mechanism to supplement their clinical experience. The modules helped to ensure educational opportunities despite the inherent variability of clinical exposure in practice. The lower scores for on-the-job training are likely reflective of fluctuating volumes and case mix that is very common in EM care spaces. Experiences with various preceptor staff and interpersonal relationships may have also been contributory to this result. These results highlighted the value of blending the onboarding model to include direct clinical experience and a structured asynchronous educational experience online.

Career Development, Reward and Recognition, and Overall Assessment domains also demonstrated positive findings. APPs rated both career management and self-assessment at 4.0, indicating that onboarding supported early professional formation. When new graduates understand expectations, receive guidance on future progression, and have opportunities to self-reflect, they experience a smoother transition to practice.^6^ Reward and Recognition results suggest that participants felt acknowledged and supported, even without incentives. This is aligned with motivational theories emphasizing mentorship, recognition, and growth opportunities as key drivers of job satisfaction in early-career providers.^4^

The Overall Assessment scores of 4.33 for teaching necessary skills and 4.0 for perceived improvement in performance further reinforced the effectiveness of the structured program. Given that new APPs previously frequently reported feeling underprepared for the transition to practice, this result signaled that structured onboarding at least improved the perception of bridging performance gaps though more research will need to be done to objectively assess if improvements were made.^4^

Qualitative findings helped reinforce the quantitative results by highlighting the role of communication, mentorship, and educational support. Participants touched on “gradual increases in expectations” which supported the importance of calibrated progression in clinical practice. The praise for the online modules highlighted the effectiveness of blended clinical and asynchronous training.

The leadership and preceptor survey findings offered support for feasibility and organizational acceptance. Leaders acknowledged that the structured onboarding model required more time and effort than the previous unstructured process, but they strongly agreed that the additional investment was worthwhile to reduce attrition. This is highly relevant in the current landscape, where APP turnover is a significant financial and operational burden for healthcare systems. The high leadership ratings for improved tracking of APP progress and the absence of undue strain on site resources further shows the feasibility and the sustainability of the process. Leadership assigned the highest rating (4.67) to improved preparedness for transition to practice, which showed that the program benefited not only participants but also team workflows and patient care quality as perceived by the preceptors. Future research could be developed to assess how comfortable supervising physicians are with APPs after this process compared to prior.

The findings demonstrated that the program was implemented successfully, with no major operational barriers and full completion of requirements by all participants. This is important given resource intensive nature of emergency medicine and the demands placed on staff during their shifts. The program showed the ability to integrate into the existing workflow with only minor scheduling adjustments which suggested that structured onboarding can be scaled across similar EM environments. High completion and satisfaction rates further demonstrated the case for expansion and ongoing research.

Despite promising results, several limitations warrant discussion. The sample size was small, a single cohort of four APPs at one clinical site. Such a limited sample will limit assumptions about generalizability and precludes inferential statistical analysis. As a feasibility study, the focus was on acceptability and perceived impact rather than performance outcomes or retention data. However, we did look for signals regarding these areas to help direct future research. Because no pre-implementation comparison group was included, the study cannot determine the magnitude of change relative to prior practice. Also, prior practice was unstructured and thus there was significant variation from new hire to new hire. Additionally, these results relied on self-report, which could introduce response bias. There may also have been bias due to low number of participants creating the concern that their responses may not have been anonymous and their hesitation to be critical of their new employer. These limitations are consistent with the goals of a feasibility study. We prioritized feasibility, acceptability, and refinement over inference.

This study provided important groundwork for future research. Longitudinal evaluation of APP performance, efficiency, and retention will provide deeper insight into the long-term impact of structured onboarding. Multi-site replication will show generalizability and allow examination of other contextual factors that may influence the program’s success. Continued use of mixed-methods approach could shed more light on how the specific components outlined above contributed to early competence and workforce stability. Cost-benefit analysis, particularly regarding turnover-related savings, may show cause for broader institutional adoption.

In summary, this feasibility study demonstrated that a structured onboarding program for new graduate APPs is practical to implement, was positively received, and aligned with organizational goals related to preparedness, engagement, and potentially retention. The program had a strong evaluation across multiple domains and consistent qualitative feedback that suggested that structured onboarding can play an important role in strengthening the early career experience of APPs in high-acuity settings. These results support continued refinement and expansion of structured onboarding strategies as healthcare systems seek sustainable approaches to transitioning the APP workforce to practice while keeping in mind cost.

## Data Availability

All data produced in the present study are available upon reasonable request to the authors

## Acknowledgements

A pre-press copy of this manuscript has been posted to the http://medRxiv.org pre-print server.

This manuscript was created as part of graduation requirements for the DMSc program of Wake Forest University School of Medicine Physician Assistant Studies.

## Supplemental Materials: Appendix A

### Week 1

- APP will meet with the Lead APP to go over expectations and the onboarding process.
- Complete all necessary medical group and site-specific orientations and trainings.
- Work 1:1 clinical shifts with Lead APP, Supervising Physician, or other designated preceptor.
- Chart reviews to occur by Supervising Physician or Lead APP.
- Complete Self-Assessment.
- Meet with Lead APP and/or Supervising Physician to go over preceptors’ feedback.
- Register for and start EM Boot Camp online modules.

### Week 2

- Work 1:1 clinical shifts with Lead APP, Supervising Physician, or other designated preceptor.
- Demonstrate competency with:
  ∘ Documentation
  ∘ Order entry
  ∘ Discharge process
  ∘ Admission process
  ∘ Evaluating Labs and diagnostic studies, assessments and plans
- Target 0.5-1.0 patients per hour (pph)
- Chart reviews to occur by Supervising Physician or Lead APP.
- Complete Self-Assessment.
- Meet with Lead APP and/or Supervising Physician to go over preceptors’ feedback.
- Continue to work on EM Boot Camp online modules.

### Week 3

- Work clinical shifts without an assigned preceptor, but as an extra clinician on the schedule.
- Staff all patients seen with Lead APP, Supervising Physician, or other designated physician.
- Demonstrate competency with:
  ∘ Evaluation and interpretation of labs and diagnostic testing
  ∘ Formation of treatment plan
  ∘ Consultation with specialists
  ∘ Presentation to attending physician Target 0.5-1.0 pph
- Chart reviews to occur by Supervising Physician or Lead APP.
- Complete Self-Assessment.
- Meet with Lead APP and/or Supervising Physician to go over shift feedback.
- Continue to work on EM Boot Camp online modules.
- 80% Documentation completion within 48 hours of the encounter.

### Week 4

- Work clinical shifts without an assigned preceptor, but as an extra clinician on the schedule.
- Staff all patients seen with Lead APP, Supervising Physician, or other designated physician.
- Demonstrate continued competency with:
  ∘ Evaluation and interpretation of labs and diagnostic testing
  ∘ Formation of treatment plan
  ∘ Consultation with specialists
  ∘ Presentation to attending physician Target ≥1.0 pph
- Chart reviews to occur by Supervising Physician or Lead APP.
- Complete Self-Assessment.
- Meet with Lead APP and Supervising Physician to go over shift feedback, this will also serve as the required North Carolina Medical Board meeting.
- 80% Documentation completion within 48 hours of the encounter.
- Complete EM Boot Camp online modules and turn in completion certificate by end of week.
- Meet with Lead APP to go over progression towards Tier 2 Compensation status, patient experience data, and expectations for transition to filling the role as a scheduled provider on the care team.
- Schedule continuing education opportunities:
  ∘ Pediatric Advanced Life Support (AHA)
  ∘ Advanced Trauma Life Support (ACS)
  ∘ Other subject or skill specific CME as identified by preceptors/self-assessment

### Week 5

- Work clinical shifts without an assigned preceptor, but as an extra clinician on the schedule.
- Staff all patients seen with Lead APP, Supervising Physician, or other designated physician.
- Demonstrate competency with:
  ∘ Evaluation and interpretation of labs and diagnostic testing
  ∘ ECG interpretation
  ∘ Formation of treatment plan
  ∘ Consultation with specialists
  ∘ Presentation to attending physician
  ∘ Procedures:
    ▪ Basic wound management
    ▪ Uncomplicated foreign body removal
    ▪ Uncomplicated laceration repair
    ▪ Uncomplicated abscess incision and drainage
    ▪ Basic fracture management Target 1.0-1.5 pph
- Chart reviews to occur by Supervising Physician or Lead APP.
- Complete Self-Assessment.
- Meet with Lead APP and/or Supervising Physician to go over shift feedback.
- 80% Documentation completion within 48 hours of the encounter.

### Week 6

- Orient to alternate care sites within the health system (Urgent Cares, stand-alone EDs, or other facilities that the APP may be asked to cover).
- Staff all patients seen with Lead APP, Supervising Physician, or other designated physician.
- Demonstrate continued competency with:
  ∘ Evaluation and interpretation of labs and diagnostic testing
  ∘ ECG interpretation
  ∘ Formation of treatment plan
  ∘ Consultation with specialists
  ∘ Presentation to attending physician
  ∘ Procedures:
    ▪ Basic wound management
    ▪ Uncomplicated foreign body removal
    ▪ Uncomplicated laceration repair
    ▪ Uncomplicated abscess incision and drainage
    ▪ Basic fracture management Target 1.0-1.5 pph
- Chart reviews to occur by Supervising Physician or Lead APP.
- Complete Self-Assessment.
- Meet with Lead APP and/or Supervising Physician to go over shift feedback.
- 80% Documentation completion within 48 hours of the encounter.

### Week 7

- Work clinical shifts without an assigned preceptor, but as an extra clinician on the schedule.
- Staff all ESI 2/3 patients seen with Lead APP, Supervising Physician, or other designated physician.
- Target independent management of ≥75% ESI 4/5 patients.
- Demonstrate continued competency with:
  ∘ Evaluation and interpretation of labs and diagnostic testing
  ∘ ECG interpretation
  ∘ Formation of treatment plan
  ∘ Consultation with specialists
  ∘ Presentation to attending physician
  ∘ Procedures:
    ▪ Basic wound management
    ▪ Uncomplicated foreign body removal
    ▪ Uncomplicated laceration repair
    ▪ Uncomplicated abscess incision and drainage
    ▪ Basic fracture management Target 1.0-1.5 pph
- Chart reviews to occur by Supervising Physician or Lead APP.
- Complete Self-Assessment.
- Meet with Lead APP and/or Supervising Physician to go over shift feedback.
- 80% Documentation completion within 48 hours of the encounter.
- Lead APP and Supervising Physician meet to discuss readiness for transition to scheduled shifts as part of the care team or need for remediation.

### Week 8

- Work clinical shifts without an assigned preceptor, but as an extra clinician on the schedule.
- Staff all ESI 2/3 patients seen with Lead APP, Supervising Physician, or other designated physician.
- Target independent management of ≥75% ESI 4/5 patients.
- Demonstrate continued competency with:
  ∘ Evaluation and interpretation of labs and diagnostic testing
  ∘ ECG interpretation
  ∘ Formation of treatment plan
  ∘ Consultation with specialists
  ∘ Presentation to attending physician
  ∘ Procedures:
    ▪ Basic wound management
    ▪ Uncomplicated foreign body removal
    ▪ Uncomplicated laceration repair
    ▪ Uncomplicated abscess incision and drainage
    ▪ Basic fracture management Target 1.0-1.5 pph
- Chart reviews to occur by Supervising Physician or Lead APP.
- Complete Self-Assessment.
- Meet with Lead APP and Supervising Physician to go over shift feedback, this will also serve as the required North Carolina Medical Board meeting.
- 80% Documentation completion within 48 hours of the encounter.
- Meet with Lead APP to go over progression towards Tier 2 Compensation status, patient experience data, and readiness for transition to filling the role as a scheduled provider on the care team.

At any time, if the APP is not meeting the above benchmarks, it will be at the discretion of the Lead APP and/or Supervising Physician to engage the APP in remediation. This can consist of informal recommendations/coaching, extension of the onboarding process, a practice improvement plan enacted and placed in the APP’s record to serve as evidence of progress towards their ability to achieve expected practice standards, or any combination thereof.

## Supplemental Materials: Appendix B

New Hire APP Survey

Questions 1-12 are 5-point scale using, Strongly Disagree, Disagree, Neutral, Agree, and Strongly Agree.

Questions 13-14 are open response.

### Domain: Career Development

#### Variable: Career Management

1. My career path is more in shape due to my participation in the training program. [18]

#### Variable: Self-Assessment

2. The self-assessments completed during the onboarding process have helped me in improving my overall required skills for transition to practice. [18]

### Domain: Training and Development

#### Variable: On the job training

3. Our organization conducts extensive training programs for its new APPs. [18]

#### Variable: Off the job training

4. The use of supplemental online modules aided in my training and preparedness for transition to practice.

### Domain: Performance Appraisal

#### Variable: Communication

5. My set targets and objective were identified by the leadership and communicated to me throughout the onboarding process. [18]

#### Variable: Periodic Review

6. I received regular formal and informal feedback in real time about my performance and progression towards the identified goals. [19]

### Domain: Reward and Recognition

#### Variable: Performance Based Rewards

7. I am more likely to achieve increased compensation tiers and a higher incentive compensation because of my participation in the onboarding process. [18]

#### Variable: Non-monetary Rewards

8. My schedule, work-life balance, and opportunity for growth are better after participating in the onboarding process.

### Domain: Employee Retention

#### Variable: Supervisor Support

9. Whenever I required extra training or coaching, I could easily approach my attending physician or a senior APP. [20]

#### Variable: Employee Engagement

10. My onboarding process motivated me to be more committed towards organizational and site-specific goals. [18]

### Overall Assessment

11. This formal onboarding process taught me, as a new employee, the skills I needed to perform my job. [18]
12. My performance improved due to the development and enhancement of skills required for transition to practice by means of the onboarding process. [18]

### Qualitative

13. What aspects of your onboarding process do you feel were most effective in preparing you for your transition to practice?
14. What improvements can be made for future APPs undergoing this onboarding process?

## Supplemental Materials: Appendix C

Preceptor/Site Leadership Survey

All questions are 5-point scale using, Strongly Disagree, Disagree, Neutral, Agree, and Strongly Agree.

1. The new structured onboarding process required more time and effort than the previous unstructured onboarding process.
2. Any additional time and effort spent on the new onboarding process was worth it to reduce attrition.
3. It was easier to track the APP’s progress through the onboarding process after the implementation of the structured onboarding process.
4. The increased onboarding time did not put undue stress on the site’s resources.
5. The new onboarding process improved the new hire APP’s preparedness for transition to practice.

## Notes

Financial Support: No financial support was used in the development of this manuscript.

Conflict of Interest: PG reports no conflict of interest. KC reports no conflict of interest. VN reports no conflict of interest.

### Competing Interest Statement

The authors have declared no competing interest.

### Funding Statement

This study did not receive any funding

### Author Declarations

Ethic committee/IRB of Wake Forest University School of Medicine gave ethical approval for this work.

